# Progressive lesion necrosis is related to increasing aphasia severity in chronic stroke

**DOI:** 10.1101/2023.06.13.23291362

**Authors:** Lisa Johnson, Roger Newman-Norlund, Alex Teghipco, Chris Rorden, Leonardo Bonilha, Julius Fridriksson

## Abstract

**Background:** Volumetric investigations of cortical damage resulting from stroke indicate that lesion size and shape continue to change even in the chronic stage of recovery. However, the potential clinical relevance of continued lesion growth has yet to be examined. In the present study, we investigated the prevalence of lesion expansion and the relationship between expansion and changes in aphasia severity in a large sample of individuals in the chronic stage of aphasia recovery.

**Methods:** Retrospective structural MRI scans from 104 stroke survivors with at least 2 observations more than 6 months apart (k=301 observations) were included. Lesion demarcation was performed using an automated lesion segmentation software and lesion volumes at each timepoint were subsequently calculated. A paired-sample t-test between initial and final scans was conducted to determine if chronic lesion volumes change over time. Finally, we investigated the association between lesion expansion and changes on the Western Aphasia Battery (WAB) in a group of participants assessed and scanned at 2 timepoints (N=54) using a GLM.

**Results:** Most participants (81%) showed evidence of lesion expansion. A t-test revealed lesion volumes at follow-up scans were significantly larger than initial scans (p<0.0001).

Change on the language performance was significantly associated with change in lesion volume (p=0.025) and age at stroke (p=0.031). The results suggest that with every 100cm^3^ increase in lesion size, language performance decreases by 8.8 points, and for every 10-year increase in age at stroke, language performance decreases by 1.9 points.

**Conclusions:** The present study confirms and extends prior reports that lesion expansion occurs well into the chronic stage of stroke. For the first time, we present evidence that expansion is predictive of longitudinal changes in language performance in individuals with aphasia. Future research should focus on the potential mechanisms that may lead to necrosis in areas surrounding the chronic stroke lesion.

## Introduction

With great advances in treatment of acute stroke in the hospital setting, the rate of stroke-related deaths has declined, leaving an ever-increasing number of stroke survivors with long-withstanding chronic deficits. Arguably the most debilitating of these deficits is *aphasia*, which refers to impairments in speech production, comprehension, or a combination of the two. The conventional understanding of chronic left hemisphere (LH) stroke-induced aphasia suggests that language recovery typically plateaus after entering the chronic stage (> 6mos post-stroke onset). However, recent evidence from our group and others has shown that about 30% of stroke survivors demonstrate marked declines over time^1–3^, even after therapeutic intervention involving intensive speech-language therapy^4,5^. Though considerable work has aimed to investigate participant factors associated with the extent of aphasia recovery, many seminal works are from small groups and case studies^6,7^ or primarily investigate the effect of aphasia treatment related to included participant factors. Research investigating the impact of detailed participant factors on aphasia recovery have suggested that much of the variance in individual recovery is dependent on both lesion- and non-lesion-related factors^8,9^.

Participant factors such as age at stroke, initial severity, presence of diabetes, and rate of exercise have been reported to be associated with both long-term recovery^1,10,11^ and treated recovery^12^. Lesion-related factors, however, have been consistently reported to explain a considerable amount of variance in individual performance^8,13–15^. In fact, lesion profiles (dependent on size and location) have been shown to explain more than 25% of the variance in long-term recovery^10^.

There is longstanding evidence to support that larger lesions result in more severe deficits^16^, however, in persons with aphasia, focal lesions can still cause significant language impairments. More recently, significant work has established the importance of spared white matter connections to minimize aphasia severity^10,17–19^. Further, in acute stroke, brain plasticity is enhanced in areas immediately surrounding the lesion (perilesional cortex)^20,21^. Similar studies have demonstrated that functional changes in perilesional cortex impact aphasia recovery even in the chronic stage^22–24^. In a follow-up study, Fridriksson et al., (2012) found that increased activation in the perilesional cortex is associated with treatment-related improvement in a language task^25^.

The pathophysiology and neuronal recovery patterns during the acute stage have been well-defined, leaving such efforts in the chronic stage unexplored. Some investigation in subacute and early chronic stroke recovery have shown evidence of necrotic tissue and edema remaining in perilesional regions which has been shown to lead to neuronal decay. In a study by Seghier and colleagues (2014), 56 patients were observed to increase in lesion volume by 1.59cm^3^ per year^26^. Further, there was a greater degree of lesion expansion for individuals whose time intervals between repeated visits was larger after controlling for baseline lesion volume, age at stroke, years post-onset, and total intracranial volume.

Therefore, given the evidence emphasizing the importance of in-tact perilesional cortex to rehabilitation and performance on speech-language tasks, gradual lesion expansion (GLE) may explain chronic behavioral declines in a sample of participants with chronic aphasia.

Additionally, and most relevant to individuals with aphasia and servicing clinicians, an individual’s rate of perilesional degeneration could also explain the inherent unreliability in treatment response to post-stroke therapy.

Therefore, the aims of the present project were two-fold. Using our large, retrospective longitudinal dataset of high-resolution structural MR images, we investigated patterns of GLE in the chronic stage of aphasia recovery. Specifically, we measured 1) longitudinal lesion volumes to investigate the presence of lesion expansion in a population with chronic aphasia and 2) the relative importance of lesion expansion in a statistical model to predict longitudinal changes in aphasia severity.

## Materials and Methods

### Participants

The present study utilized retrospectively collected longitudinal data of individuals in the chronic stage of aphasia recovery (≥ 6 months post-stroke at initial scan and testing timepoint) drawn from the open-source Aphasia Recovery Cohort (ARC) dataset. Upon initial scan, participants (or their caretaker) completed a detailed case history survey that inquired about detailed demographic and health data. A total of 104 participants (33 female) with at least two imaging sessions (at least 1 month apart) met inclusion criteria available resulting in a total of 303 imaging observations across all participants. Data from two scanning sessions were removed from the analysis due to presence of significant imaging artifacts that hindered automated lesion detection methods (described below). Therefore, a total of 301 scans for 104 participants were included in the present study.

### Scanning Protocol

Magnetic resonance imaging (MRI) scans at each time point were collected for an independent study or at baseline for a treatment study at the Center for the Study of Aphasia Recovery (C-STAR) laboratory at the University of South Carolina or the Medical University of South Carolina. For the purposes of the present study, only structural MRI images (T1s) were examined. Lesions were automatically identified and delineated using freely available lesion-segmentation software (described below), and lesion volumes were calculated based on the output of this software. High-resolution T1 images were acquired using a Siemens Trio/Prisma 3T scanner equipped with a 20-element head-neck coil with the following parameters: an MP-RAGE sequence with 1mm isotropic voxels, a 256×256 matrix size, a 9-degree flip angle, and a 92-slice sequence with repetition time = 2250 ms, inversion time = 925ms, and echo time = 4.11ms.

### Lesion Segmentation Protocol

Lesion segmentation was conducted using Lesion Identification with Neighborhood Data Analysis (*LINDA*), an automated lesion segmentation software^27^, with default settings, resulting in a single probabilistic lesion mask for each participant at each time point (i.e., 301 total masks). Subsequently, each probabilistic lesion mask was reviewed by authors AT and/or LJ to ensure successful lesion demarcation. Upon review, it was noted that scans with considerable artifacts (caused by excessive motion or lesions that interfere with the lateral ventricles) resulted in poor automated lesion demarcation. Therefore, these scans were reviewed by trained lesion raters and lesions were redrawn manually. An additional two scans were removed from the dataset due to scan quality being affected by excessive motion. Following automatic lesion segmentation, the final prediction mask output from LINDA (Prediction3) in native space was used to calculate the total lesioned voxels (cc) for all participants’ scans.

Change in lesion volume was calculated by subtracting volume at initial scan from lesion volume at the follow-up scan with positive numbers indicating greater expansion. Change in lesion volume was used as the primary variable of interest in the analyses described below.

### Evaluating Longitudinal Lesion Expansion

A total of 104 participants (33 female; k=301 observations) who had at least two scanning sessions (M=2.89, SD=0.93) were included in an analysis to evaluate longitudinal lesion expansion. A paired *t*-test was conducted for lesion volumes at initial and final follow-up scans to determine if there was a significant difference in lesion volume between scans. A significant difference was considered if *p*-value < 0.05.

### Lesion Expansion as a Predictor of Longitudinal Behavior

To investigate the impact of lesion expansion on behavior, a general linear model (GLM) was created for a subset of participants who had longitudinal behavioral and imaging data available (N=54). Participants were included in the present analysis if they were assessed using a standardized aphasia severity battery, the Western Aphasia Battery-Revised (WAB-R)^28^, twice at least 3-months between assessments (M=38.5; SD=27 months) and received a high-resolution MRI scan within 10-days of the WAB-R assessment. It is important to note that the WAB-R was never used as an outcome measure in any treatment study, therefore the Aphasia Quotient score from the WAB-R (WAB AQ) was a direct reflection of treatment gains.

Independent variables included in the GLM were based on prior studies^1,8^ which found significant relationships between the factors and the dependent variable, change in WAB AQ between assessments. These variables included: initial severity (NIH Stroke Scale; NIHSS), age at stroke, and days between assessments. A measure of lesion expansion (lesion volume at follow-up–lesion volume at initial scan) was the dependent variable of interest. Relationships between independent variables and WAB AQ change were considered significant if *p*-values < 0.05.

## Results

### Evaluating Longitudinal Lesion Changes

The mean time post stroke onset at the time of initial scan was 3.4 years (range = 6 mos–17.3 yrs), which is well within the range of what is considered the chronic stage of stroke recovery. Table 1 presents demographic information for participants included in the investigation of longitudinal expansion.

**Table 1:**
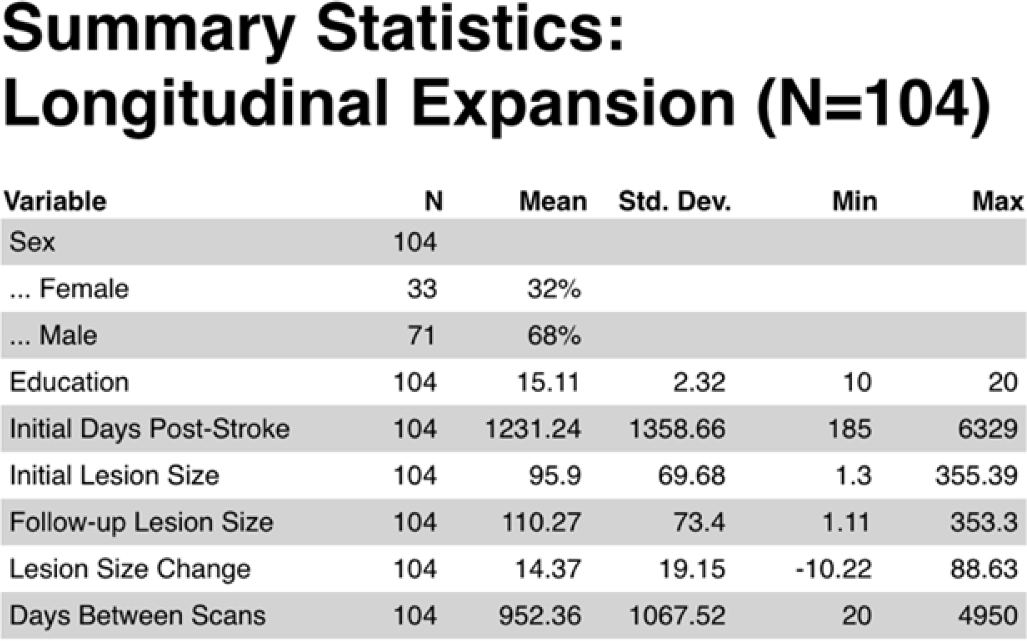
Descriptive statistics for cohort with at least 2 structural images used to evaluate GLE. Both GLE and Days Between Scans were calculated using data from the final scan and the initial scan for each participant. Lesion sizes are measured in cubic centimeters (cc).

To better visualize GLE, Figure 1 demonstrates expansion across the group depending on percent of lesion overlap between initial and final scans. In this figure, individuals are binned depending on the percent of lesion overlap, where those with the most lesion overlap (i.e., demonstrated the most lesion expansion) are shown in yellow and those with the least lesion overlap (i.e., the least lesion expansion) are shown in gray. Across the group, the insula was the region most often damaged due to the stroke lesion and average initial lesion mask for each binned group and the average follow-up lesion mask is shown.

**Figure 1:**
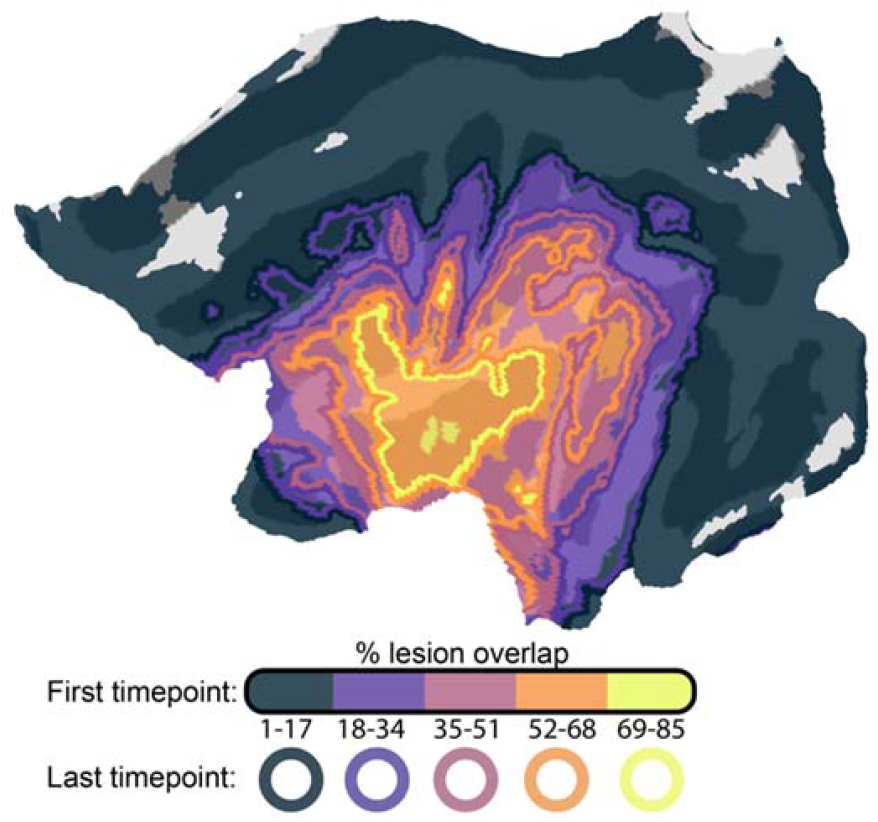
Flat map depicting lesion expansion for the group (N=104). Participants were binned based on percent lesion overlap between initial and follow-up scans (greater overlap indicates greater lesion expansion). Solid-filled regions indicate the lesioned areas of greatest overlap in that binned group at initial scan, and the ring of the same color indicates the average lesion border at the last (follow-up) scan.

Figure 2 presents the lesion volumes at each scanning timepoint (collected at least 1-month between scans) available for each participant (indicated by circle color) in the chronic stage of stroke recovery. The slope of each participant’s lesion size trajectory is shown to detail participants with expanding lesions (red line) or participants with relatively stable lesion size (black line). For ease of visualization and interpretation of Figure 2A, years post-stroke at time of scan (x-axis) was log transformed. Overall, the average change in lesion size was 14.4cc (SD=19.2cc; range=-10.2cc–88.6cc), showing that the majority of participants’ chronic lesions (85/104, 81%) expanded over time. To demonstrate that this effect is present even years beyond stroke occurrence, Figure 2B presents the lesion volumes at each scanning timepoint with participants binned by the time post-stroke at the initial scanning timepoint.

**Figure 2:**
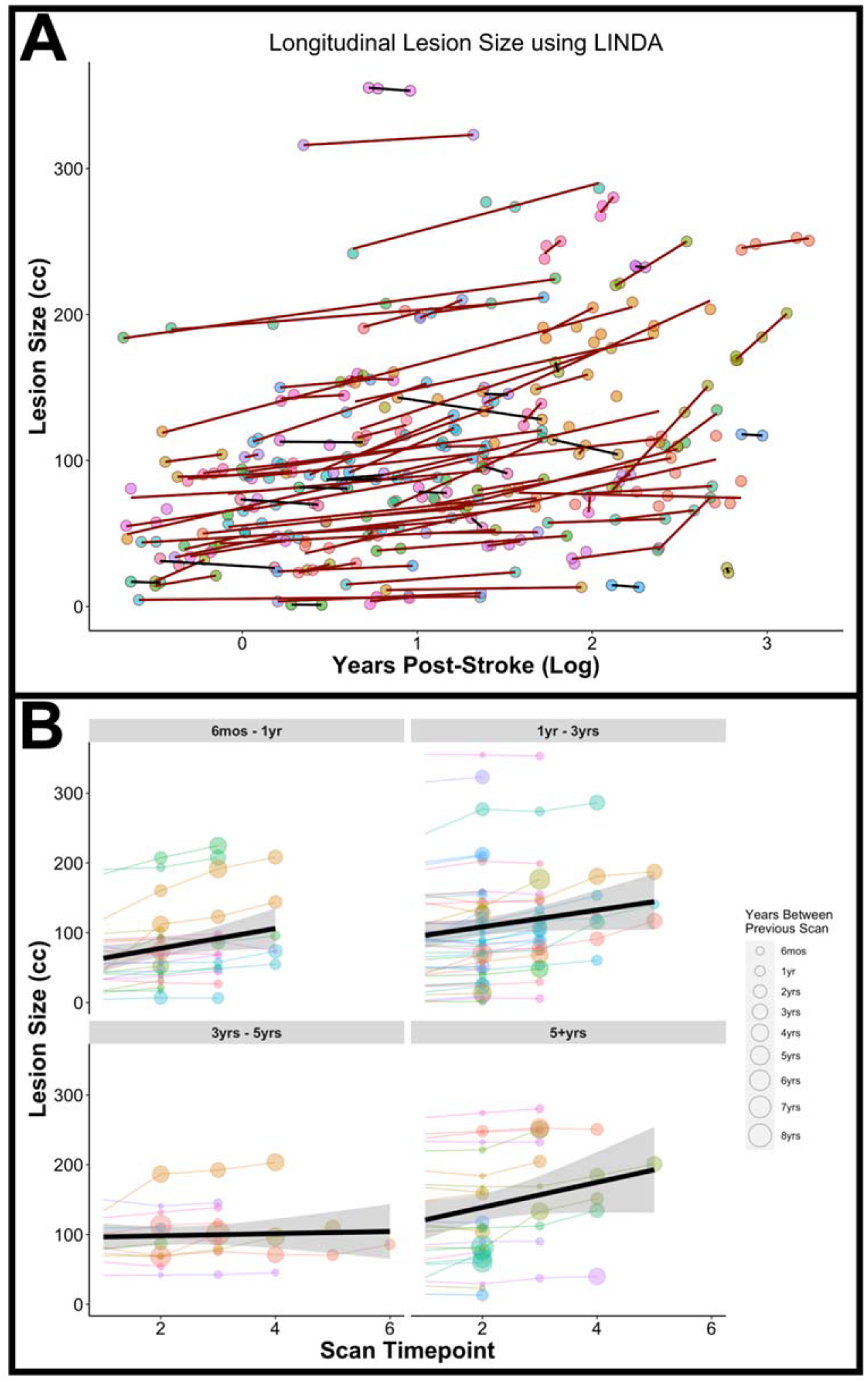
(A) Change in lesion volume over time for 104 participants with chronic LH stroke and aphasia. Red lines indicate lesions that have increased in size between initial scan and final follow-up scan (positive slope); black lines indicate lesions that have remained stable over time (negative slope); (B) Change in lesion volume for participants grouped by the time post-stroke at initial scan. Size of the participant node indicates time between scanning sessions.

A two-tailed paired-samples *t*-test revealed that lesion sizes at the final follow-up scans were significantly larger (M=110.3cc, SD=73.4cc) compared to lesions at initial scans (M=95.9cc, SD=69.7cc); t(103)=-7.7, p<0.0001 (Figure 3).

**Figure 3:**
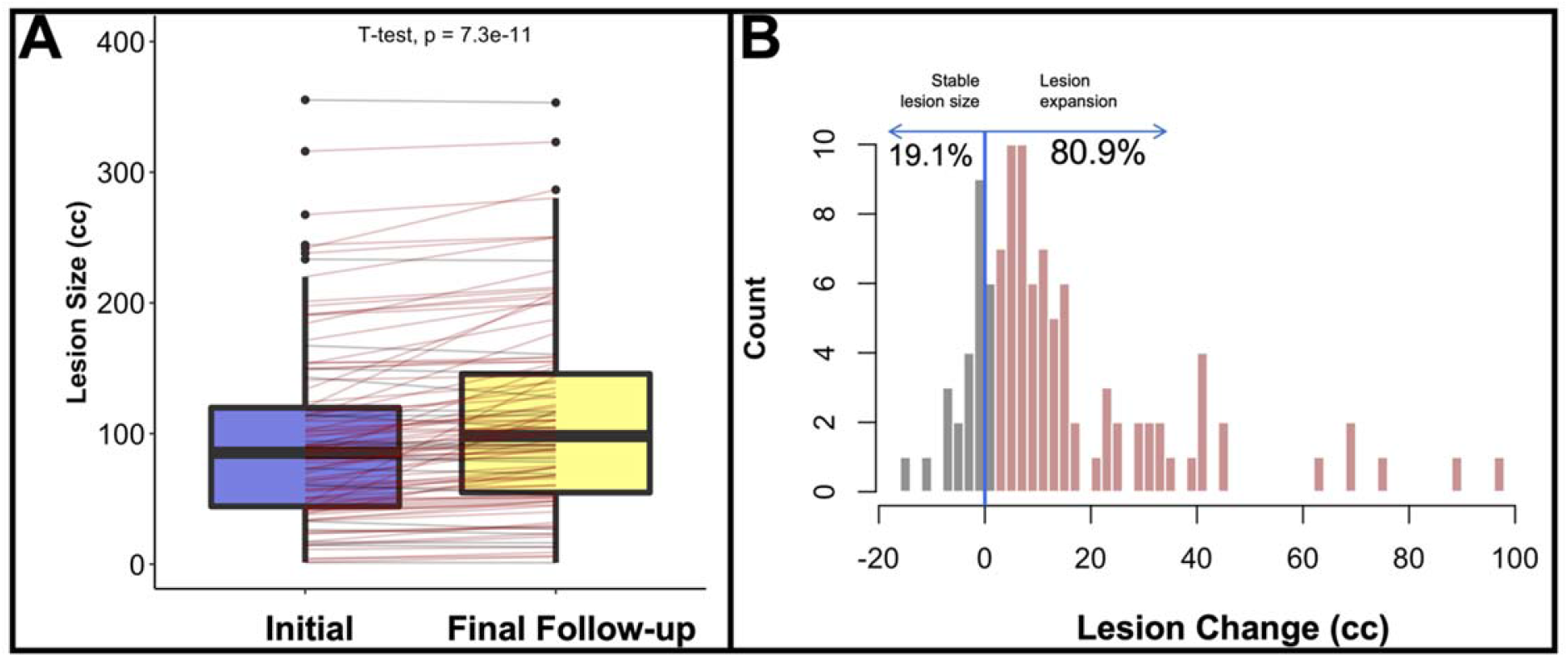
(A) Boxplot depicting lesion size at initial and final follow-up scans. A paired one-sample t-test indicates lesion volumes were significantly larger at final follow-up (p<0.0001); (B) Histogram demonstrating proportion of expanders and non-expanders in the sample (N=104).

### Effect of Longitudinal Lesion Changes on Behavior

Table 2 presents demographic information for participants included in the analysis investigating how change in lesion volume may explain longitudinal WAB AQ change.

**Table 2:** Descriptive statistics for subsample with both baseline and follow-up WAB AQ assessments as well as both baseline and follow up MRI examinations WAB = Western Aphasia Battery, AQ = Aphasia Quotient, NIHSS = National Institutes of Health Stroke Scale, Lesion sizes are measured in cubic centimeters (cc).

| Variable | N | Mean | Std. Dev. | Min | Max |
| --- | --- | --- | --- | --- | --- |
| Sex | 54 |  |  |  |  |
| ... Female | 14 | 26% |  |  |  |
| ... Male | 40 | 74% |  |  |  |
| Education | 54 | 15.31 | 2.38 | 10 | 20 |
| Initial Days Post-Stroke | 54 | 1228.35 | 1168.77 | 185 | 6127 |
| Initial Lesion Size | 54 | 105.17 | 58.75 | 4.44 | 277.03 |
| Age at Stroke | 54 | 54.44 | 10.84 | 31 | 75 |
| Initial NIHSS | 54 | 5.59 | 3.42 | 1 | 14 |
| Lesion Size Change | 54 | 21.55 | 24.83 | -10.22 | 118.58 |
| Days Between Scans | 54 | 1131.28 | 870.51 | 88 | 4539 |
| WAB AQ Change | 54 | 2.07 | 7.58 | -22.7 | 19.5 |

**Table 3:** Results from GLM predicting change in WAB AQ performance. Bolded p-values indicate significant (p<0.05) predictors.

| Predictors | Estimates | CI | p |
| --- | --- | --- | --- |
| (Intercept) | 13.102 | 2.179 – 24.026 | **0.020** |
| Lesion Size Change | -0.088 | -0.165 – -0.012 | **0.025** |
| Initial NIHSS | 0.323 | -0.193 – 0.839 | 0.215 |
| Age at Stroke | -0.188 | -0.358 – -0.018 | **0.031** |
| Days Between WABs | -0.002 | -0.004 – 0.001 | 0.147 |
| Observations | 54 |  |  |
| R<sup>2</sup> / R<sup>2</sup> adjusted | 0.238 / 0.175 |  |  |

**Predicting Change in WAB AQ**
| *Predictors* | *Estimates* | *CI* | *p* |
| --- | --- | --- | --- |
| (Intercept) | 13.102 | 2.179 – 24.026 | **0.020** |
| Lesion Size Change | -0.088 | -0.165 – -0.012 | **0.025** |
| Initial NIHSS | 0.323 | -0.193 – 0.839 | 0.215 |
| Age at Stroke | -0.188 | -0.358 – -0.018 | **0.031** |
| Days Between WABs | -0.002 | -0.004 – 0.001 | 0.147 |
| Observations | 54 |  |  |
| R<sup>2</sup> / R<sup>2</sup> adjusted | 0.238 / 0.175 |  |  |

A GLM was conducted to investigate the effect of change in lesion volume on long-term behavioral performance in 54 participants. We observed a main effect of age at stroke on change in WAB AQ (*p*=0.031), as well as a main effect of change in lesion volume on change in WAB AQ (*p*=0.025). Results from the model demonstrate that every 100cc of lesion expansion was associated with 8.8 points decrease on the WAB AQ, and for every 10-year increase at age of stroke, WAB AQ decreased by 1.9 points. Figures 4 and 5 present the main effects between lesion size change and age at stroke on WAB AQ, respectively. Change in WAB AQ performance between assessments was not associated with time between assessments or initial stroke severity.

**Figure 4.**
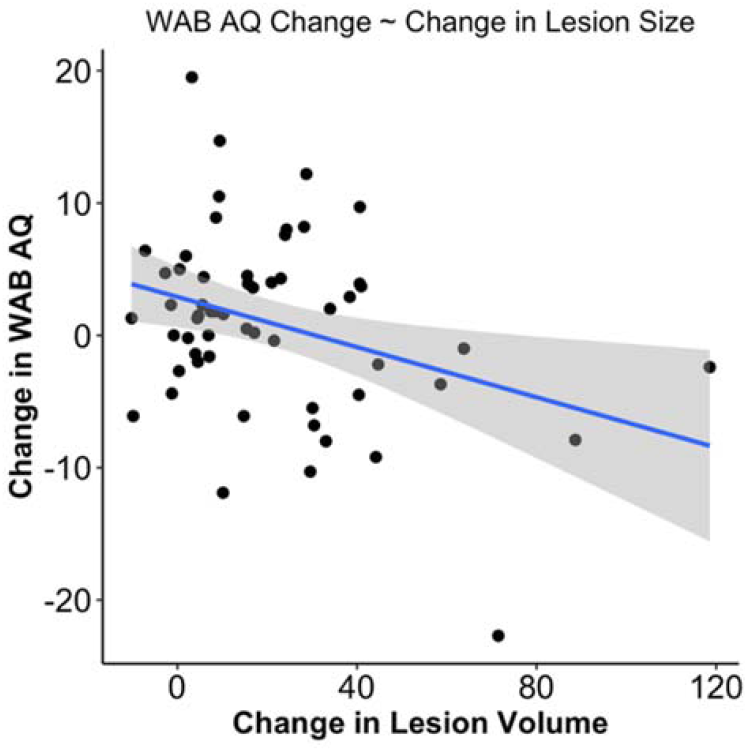
(left): Scatterplot demonstrating the significant main effect of change in lesion volume (cc) as a predictor of change in WAB AQ.

**Figure 5.**
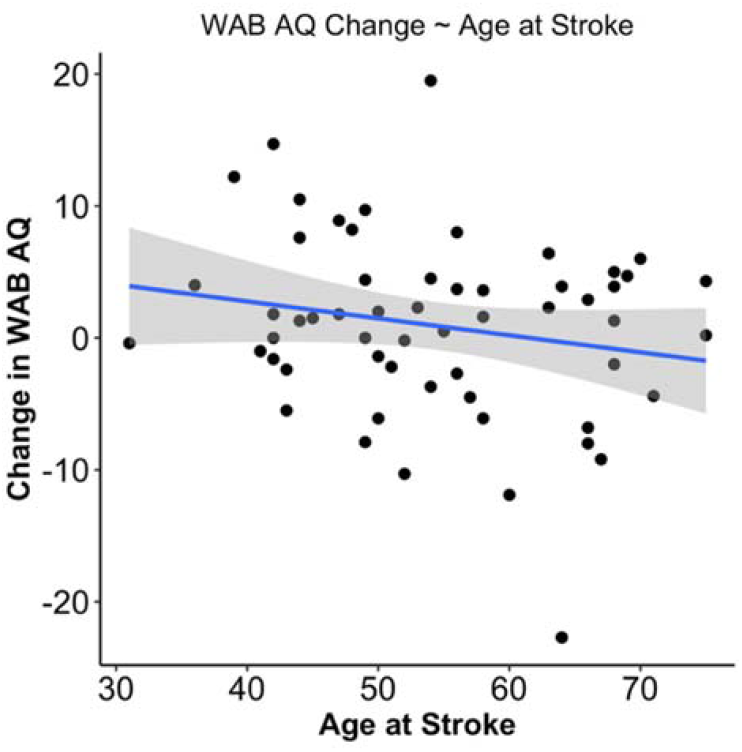
(right): Scatterplot demonstrating the significant main effect of age at stroke as a predictor of change in WAB AQ.

## Discussion

Previous research indicates that cortical damage associated with acute stroke, as measured by lesion size, continues to evolve with time and that this increase is due to both accelerated global atrophy and, more targeted decreases in cortical integrity at perilesional sites^26,29^. However, the relationship between GLE and behavioral sequalae of stroke remains wholly unexplored. The current study addressed this gap by examining the relationship between GLE and language impairment in a large cohort of individuals with LH stroke and aphasia.

Consistent with a study by Seghier and colleagues (2014) that examined GLE, we report that lesion expansion was present in the chronic timeframe in the majority of participants, and we extend these results, for the first time, to individuals with chronic LH stroke and aphasia. Direct comparison of lesion size at two time points (mean separation between time points = 31 months) confirmed that lesion volumes were significantly larger at follow-up testing. One important contribution of the present study is the investigation of long-term GLE in individuals years after their stroke event (minimum 6 months post-stroke at initial scan). Seghier and colleagues’ paper included participants within the late subacute stages (≥3 months post-stroke), when stroke-related edema could still be present. Therefore, the present study not only validates the findings that lesion expansion is present in individuals beyond the acute stage but also provides evidence that individuals years post-stroke demonstrate GLE as well.

Finally, the impact of GLE on behavioral change was investigated in a subset of participants who had both MRI scans and WAB assessments available at both initial and follow-up assessments. A general linear model, with change in WAB AQ as the dependent variable, revealed that both change in lesion volume and age at stroke were significantly associated with change in WAB AQ (p=0.025; p=0.031, respectively). This is the first study, to our knowledge, to demonstrate a relationship between GLE and behavioral change in a sample of individuals with chronic stroke aphasia.

## Pathophysiology of Stroke Lesions

While the pathophysiology of and neuronal recovery patterns associated with the acute stage of stroke have been well-defined^30–32^, explanations of analogous changes in the chronic stage of recovery remain largely unexplored. One potential explanation for perilesional degradation observed in the current study is that GLE reflects persistent local Wallerian degeneration of short-range intracortical connections driven by lack of input from areas immediately surrounding the stroke lesion. Though typically discussed in terms of its impact on the acute to sub-acute recovery stages, there is evidence to suggest WD continues in some instances through the chronic stage of stroke recovery. For example, Werring and colleagues confirmed that degeneration can be observed in stroke patients 2-6 months post infarct, although it was noted that extent of WD was variable across individuals^33^. Interestingly, there is evidence that other types of MRI scans, most notably diffusion tensor imaging (DTI), which measures white matter tract integrity, can be used as an alternative measure of changes due to WD, a fact which opens the interesting possibility of examining DTI in future chronic stroke studies^34^.

An important consideration in the current study is the fact that our patients were specifically chosen for study inclusion based on the presence of aphasia. Aphasia typically results from strokes that damage brain sites in the putative left hemisphere language system. Damage to these areas is most often the result of ischemia or hemorrhage of the middle cerebral artery (MCA), which is situated between two watershed areas, areas that exist at the border between two major cerebral arteries. One hypothesis for observed lesion expansion is that watershed areas are more susceptible to the neural degeneration observed in GLE^29^. This may be due to the proximity between two adjacent arteries, causing watershed zones to particularly susceptible to alterations of hemodynamic conditions^35^, which can result in hypoperfusion, eventually resulting in neural degeneration^36^. In animal models, chronic hypoperfusion has been shown to contribute to neuronal death, which can lead to expanding lesions in these areas post-stroke. Kudo and colleagues demonstrated that gerbil brains post-stroke had considerable hypoperfusion in watershed zones surrounding a stroke lesion, and all lesions expanded after 12 weeks, particularly in the middle frontal gyrus, which is adjacent to the inferior frontal gyrus that, when damaged, is associated with speech production deficits^37^. This reduced hypoperfusion can appear intact but can create what is described as a “functional lesion,” where neurons are viable but are not functioning properly^38–40^. Indeed, evidence from Thompson and colleagues (2017) demonstrates perilesional hypoperfusion in chronic stroke lesions (0-6mm surrounding the lesion), which is associated with behavioral scores in persons with aphasia^38^. Future studies might examine GLE and its relationship with behavioral improvement in alternative stroke populations in which watershed areas are not so directly implicated.

## Gradual Lesion Expansion Predicts Change in Behavioral Performance

Language performance in the chronic stage of aphasia is dynamic with nearly 30% of individuals with demonstrating significant gradual decline in linguistic performance, even after receiving intensive speech-language therapy. Measures of neural health have recently garnered attention in the stroke-aphasia literature due to their association with chronic behavioral declines in spared contralateral regions^10,11,41^. Specifically, the presence and extent of markers of small vessel disease (i.e., white matter hyperintensities, lacunes, enlarged peri-vascular spaces) in the contralateral hemisphere have been proposed as a measure of overall brain health, which is significantly related to longitudinal performance. Evaluating the relative health of the ipsilateral hemisphere, however, poses a challenge due to the hyperintense tissues surrounding the lesion proper, and the often minimal, spared tissue that is able to be evaluated if a lesion is particularly large.

Our understanding of the relationship between brain health and behavioral outcomes in the ipsilesional hemisphere is less clearly developed. For example, one study found that fMRI changes in perilesional areas was not associated with behavioral change. Another found that cerebral blood flow in perilesional areas was similarly unrelated to improvement. In contrast, early evidence from our group^25^ and others have demonstrated the importance of perilesional integrity to predict overall severity and aphasia rehabilitation^42^. If we assume that perilesional tissue degraded most in individuals with greater GLE, then the current study can be interpreted as providing further support for the idea that health of perilesional tissue is critical to long-term recovery. Such an interpretation has clear implications for therapeutic interventions designed to prevent/slow brain atrophy in stroke survivors, highlighting the need to develop and test interventions directed at perilesional tissues.

## Limitations and Future Directions

Though the present study presents evidence that perilesional necrosis is associated with long-term behavioral change in chronic stroke aphasia, there remains to be determined the cause of the underlying mechanism that triggers lesion expansion. One possibility could be due to genetic or health factors that could result in necrosis of the tissue(s) surrounding the lesion. Future studies should examine the relationship between comorbidities (including small vessel disease, diabetes, and activity level) and changes in aphasia severity as these associated health factors have been shown to influence overall aphasia severity and recovery. Unfortunately, these data were not available for all participants with longitudinal imaging and behavioral assessments in the current.

We were limited in the longitudinal behavioral data available for the cohort of participants with longitudinal imaging. The present results provide evidence that GLE is related to changes in performance on a language assessment, however, it is unclear if other behavioral tasks (i.e., measures of cognition) also demonstrate a similar association. Prior studies suggest observed GLE could be an early marker of vascular dementia^26^, therefore, future studies should investigate the impact of GLE on more global measures of cognition.

Although it does not affect the interpretation of the current findings, it would be interesting to examine the impact of acute treatment (i.e., via pharmacological intervention and/or tissue plasminogen activator, and timeliness of administration in the hospital setting) impacts GLE. Given the nature of recruiting participants in the chronic stage of recovery, we were unable to investigate these factors, but future longitudinal studies could shed light on a potential mechanism that predicts GLE in chronic recovery.

## Conclusions

Conventional wisdom regarding chronic stroke lesions suggests that lesion borders do not change past the acute and subacute stages of injury. The results from the present study provide compelling evidence for GLE well into the chronic stages of left hemisphere MCA stroke-induced aphasia. Moreover, we show, for the first time, GLE is relevant to disease prognosis and potentially treatment response. GLE provides another mechanistic explanation for declines observed in chronic stroke recovery and these findings may have important implications for researchers and clinicians interested in maximizing performance and recovery by minimizing/slowing the evolution of post-insult damage to the brain.

## Data Availability

The data used in this manuscript is publicly available on OpenNeuro.

https://openneuro.org/datasets/ds004512

## Disclosures

The authors have no financial conflicts of interest.

## Notes

### Competing Interest Statement

The authors have declared no competing interest.

### Funding Statement

This work was supported by the National Institute on Deafness and Other Communication Disorders (Fridriksson: R03 DC005915, R01 DC008355, R01 DC009571, R03 DC010262, R01 DC011739, R21 DC014170, P50 DC014664; Basilakos: T32 DC014435).

### Author Declarations

Each participant included in the study provided written informed consent for participation in accordance with the Declaration of Helsinki.

